# Association of Non-High-Density Lipoprotein Cholesterol to High-Density Lipoprotein Cholesterol Ratio (NHHR) with Cardiovascular-Kidney-Metabolic (CKM) Syndrome Stages: Evidence from a Nationally Representative U.S. Cohort (NHANES 2005–2018)

**DOI:** 10.1101/2025.09.21.25336290

**Authors:** Jiahui Qin, Jiaxin Tan, Lifeng Huang, Jian Xie

## Abstract

**Background:** The Non-HDL-C/HDL-C ratio (NHHR) is an emerging lipid marker, but its role in Cardiovascular-Kidney-Metabolic (CKM) syndrome remains unclear.

**Objective:** This study investigated NHHR’s association with CKM stages in a U.S. cohort.

**Methods:** A total of 14638 participants were included out of 51,199 from the NHANES 2005-2018 survey. Analyzed using a survey weighting approach to ensure a nationally representative sample. The connection of NHHR with CKM syndrome was assessed via multivariate logistic regression, restricted cubic splines (RCS), and subgroup analyses. The predictive performance of NHHR across different CKM stages was assessed via the Receiver operating characteristic (ROC) curve.

**Results:** The NHHR level in patients with CKM syndrome was higher than in patients without (2.87±1.14 vs 2.35±0.93 mg/dL, P<0.001). The multivariate logistic regression analysis suggested a positive connection between NHHR and CKM syndrome. The RCS analysis revealed a nonlinear association between the NHHR and CKM syndrome (nonlinear p < 0.001). ROC curve analysis demonstrated that NHHR had the best discriminatory ability for CKM stage 2 (area under the curve 0.673). Subgroup analyses revealed that age, race, and education significantly influenced the connection between NHHR and CKM syndrome.

**Conclusion:** This research confirmed that NHHR was positively correlated with CKM syndrome and had the best predictive ability for CKM stage 2, suggesting that NHHR may assist in the early diagnosis and intervention of CKM syndrome.

## Introduction

Cardiovascular–kidney–metabolic syndrome (CKM) is a novel spectrum of diseases that has garnered increasing attention in recent years and involves complex interrelationships among metabolic syndrome (MetS), cardiovascular disease (CVD), and chronic kidney disease (CKD)[1]. The clinical manifestations of CKM syndrome progress from a single metabolic abnormality in the early stages to simultaneous heart and kidney failure in the late stages[2]. Notably, in the advanced stages of CKM, patients are often affected by diabetes, renal failure, and a heightened risk of cardiovascular events, further exacerbating the disease burden and placing immense pressure on both individual health and public healthcare systems[3]. Therefore, identifying effective biomarkers for early surveillance and management is crucial.

The ratio of non-high-density lipoprotein cholesterol (non-HDL-C) to high-density lipoprotein cholesterol (HDL-C) (NHHR) is an emerging lipid metric that has been shown to have an important prognostic role in many cardiovascular diseases, particularly in assessing the risk of atherosclerosis[4–6]. In recent years, multiple studies have further revealed the critical role of NHHR in metabolic disorders and chronic kidney disease. For instance, Kim et al. found that NHHR has excellent predictive power in metabolic syndrome, exceeding the predictive value of apoB/apoA1 ratio[7]. Furthermore, Wen et al. also confirmed that NHHR is closely related to the prevalence of CKD[8]. These diseases are closely associated with CKM syndrome, indicating that NHHR may have a potential role in CKM syndrome. As a comprehensive lipid index, NHHR can reflect the imbalance of lipid metabolism, and lipid metabolism disorder plays a key role in developing CKM syndrome[1,9]. Therefore, studying the relationship between NHHR and CKM syndrome may provide more effective early detection and intervention indicators.

Thus, this study aims to systematically analyze the changing characteristics of NHHR in different stages of CKM syndrome and uses a large sample, multi-factor analysis method to comprehensively evaluate the association between NHHR and CKM syndrome, providing a new perspective and theoretical basis for the study of CKM syndrome.

## Methods

Data for this research were obtained from the National Health and Nutrition Examination Survey (NHANES) in the United States from 2005 to 2018. The survey uses stratified random sampling to collect data every 2 years, covering a wide range of health and nutrition information. However, due to factors such as the survey method and sample selection, the data may have certain limitations, such as insufficient representation of some special populations. The NHANES research was approved by the Ethics Review Board of the National Center for Health Statistics, and each participant provided written informed consent to ensure ethical compliance. For detailed participant information, refer to NHANES website (https://www.cdc.gov/nchs/nhanes/index.html).

### Research population

This study initially included 51,199 survey participants from the NHANES database from 2005-2018. After screening, 36,561 participants under the age of 20 and those lacking data on gender, age, education, race, poverty-to-income ratio (PIR), smoking, CKM syndrome, and NHHR were excluded. For missing data, the multiple imputation method was used. During the screening process, statistics showed that the proportion of missing data was less than 5%, which had little impact on the research results. As a result, 14,638 eligible participants were encompassed in the study. The sample selection process is displayed in **Figure 1**.

**Figure 1.**
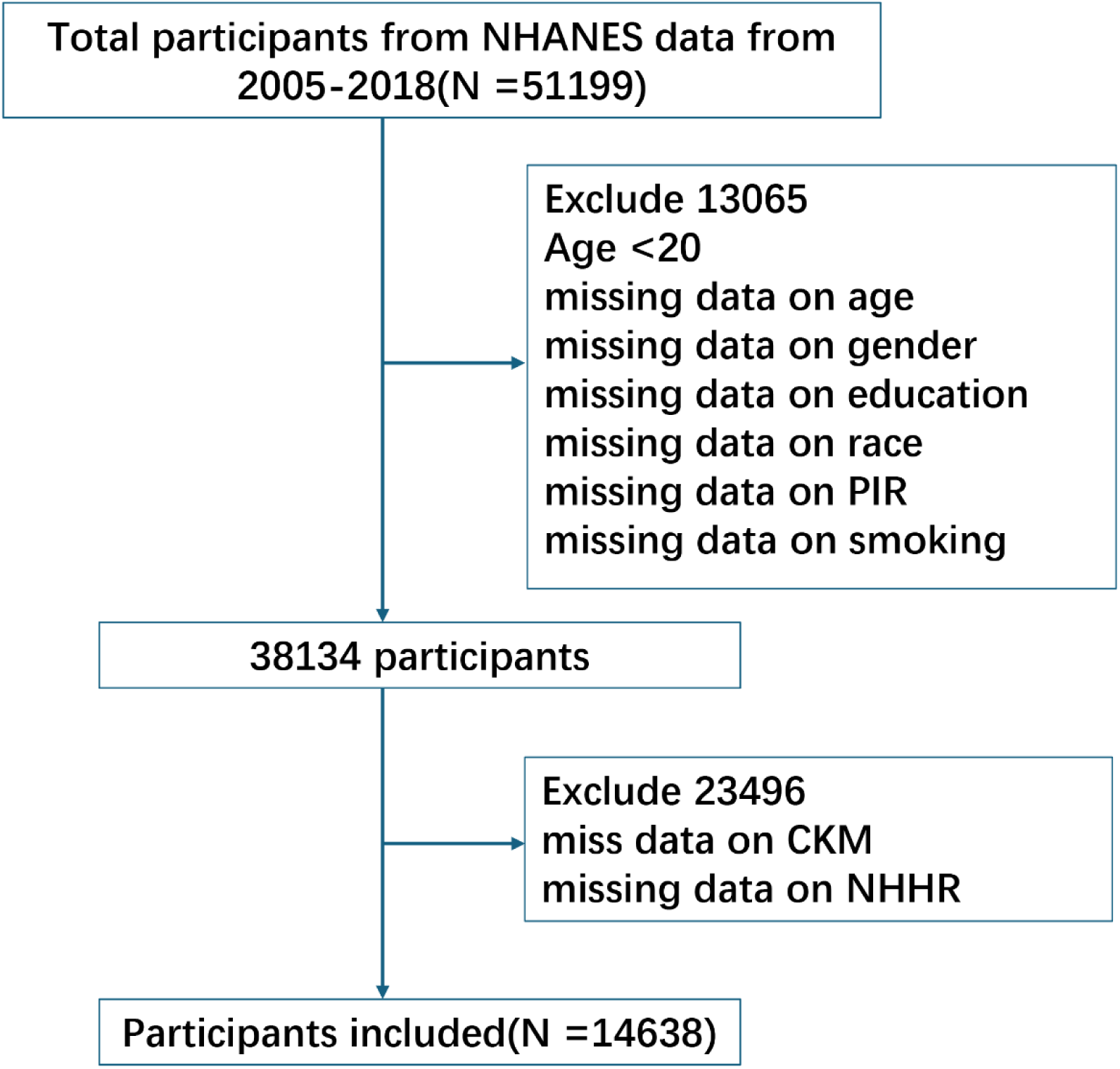
NHANES participant selection flowchart.

### Ascertainment of CKM stages

Built upon the 2023 American Heart Association Presidential Advisory on CKM Health[10], CKM syndrome is divided into stages 0 to 4. Each stage is defined as follows: CKM stage 0 features the absence of any CKM risk factors (normal BMI, waist circumference, blood glucose, blood pressure, lipids, and the absence of CKD or CVD); CKM stage 1 is characterized by early-stage metabolic abnormalities (BMI ≥25 kg/m² or ≥23 kg/m² in Asians, increased waist circumference, or prediabetes); CKM stage 2 is characterized by having metabolic risk factors (e.g., hypertriglyceridemia, hypertension, diabetes, or metabolic syndrome) or intermediate- to high-risk CKD; CKM stage 3 is characterized by having very-high-risk CKD or a high predicted 10-year risk of CVD (≥20%); CKM stage 4 is characterized by a diagnosis of CVD (e.g., coronary artery disease, heart failure, stroke, etc.). **Table S1** provides a detailed description of the definitions of each stage.

### NHHR assessment

In this research, NHHR served as the independent variable. The final NHHR data for the participants were computed by deducting the HDL-C level from the total cholesterol (TC) level and dividing the result by the HDL-C level. The laboratory test results from the NHANES database were leveraged to calculate the total cholesterol and HDL-C levels.

### Demographic characteristics and other covariates

Several covariates, including age, gender, race, educational level, PIR, and smoking, were included in our study to control for potential confounders that related to NHHR and CKM syndrome. Mexican American, non-Hispanic Black, non-Hispanic White, other Hispanic, and other race were the categories for race and ethnicity. Less than 9th grade, 9th–11th grade, high school graduate, some college or AA degree, and college graduate or beyond were the categories for level of education. Smoking status was identified by inquiring participants whether they had consumed at least 100 cigarettes throughout their lifetime, with response options of yes or no. Regarding the family income-to-poverty line ratio, PIR was split into two groups: less than 2 and more than or equal to 2.

### Statistical analysis

To ensure the selected samples were representative of the U.S. population, we employed stratification, clustering, and sample weighting in our analysis of NHANES data. Stratification was applied based on age, gender, and other demographic factors to control for potential confounding variables. Clustering was used to account for the inherent clustering of the sample, thereby reducing estimation errors. Sample weighting was performed to align the sample distribution with the U.S. population demographics (age, gender, and race), enhancing the generalizability of our findings. Continuous variables are presented as means ± standard deviation (SD). Differences in continuous variables across NHHR quartiles were assessed using weighted one-way analysis of variance(ANOVA), while comparisons between groups with CKM syndrome and without CKM syndrome were performed using Wilcoxon rank-sum test. Categorical variables are expressed as numerical values (percentages) and analyzed using the chi-square test. Multivariate logistic regression was used to evaluate the association between NHHR and CKM syndrome, and three different models were constructed to gradually adjust for age, gender, socioeconomic and lifestyle factors to analyze their independent effects. Model 1 did not include any covariate adjustments. Model 2 was adjusted for age and gender. Model 3 was further adjusted for age, gender, race, PIR, education, and smoking. Additionally, restricted cubic spline (RCS) analyses were utilized to explore potential nonlinear relationships between NHHR and CKM syndrome, visually representing dose-response relationships. Odds ratio (OR) and 95% confidence intervals (CI) were calculated for NHHR quartiles, with the lowest quartile serving as the reference group. To assess the predictive performance of NHHR for CKM syndrome stages 1–4, we calculated receiver operating characteristic (ROC) curves and the area under the curve (AUC). Subgroup analyses were conducted to examine potential effect modifications by gender, race, education, PIR, and smoking on the association between NHHR and CKM syndrome. All statistical analyses were performed using RStudio 4.2.1 and SPSS 26.0.

## Results

### Baseline characteristics of the research participants stratified by CKM syndrome

**Table 1** displays the baseline characteristics of the participants stratified by CKM syndrome. 14638 participants were included in the research; the mean age of the participants was 47.65 ± 16.70 years, 45.34% (n=6637) of them were females, and they were categorized into participants with CKM syndrome (9284) and those without CKM syndrome (5354). The participants with CKM syndrome were predominantly male. Notably, patients with CKM syndrome had significantly higher NHHR levels than those without (2.35±0.93 vs. 2.87±1.14 mg/dL, P<0.001). Remarkable differences in age, race, education, PIR, and smoking status were found between the two groups.

**Table 1.**
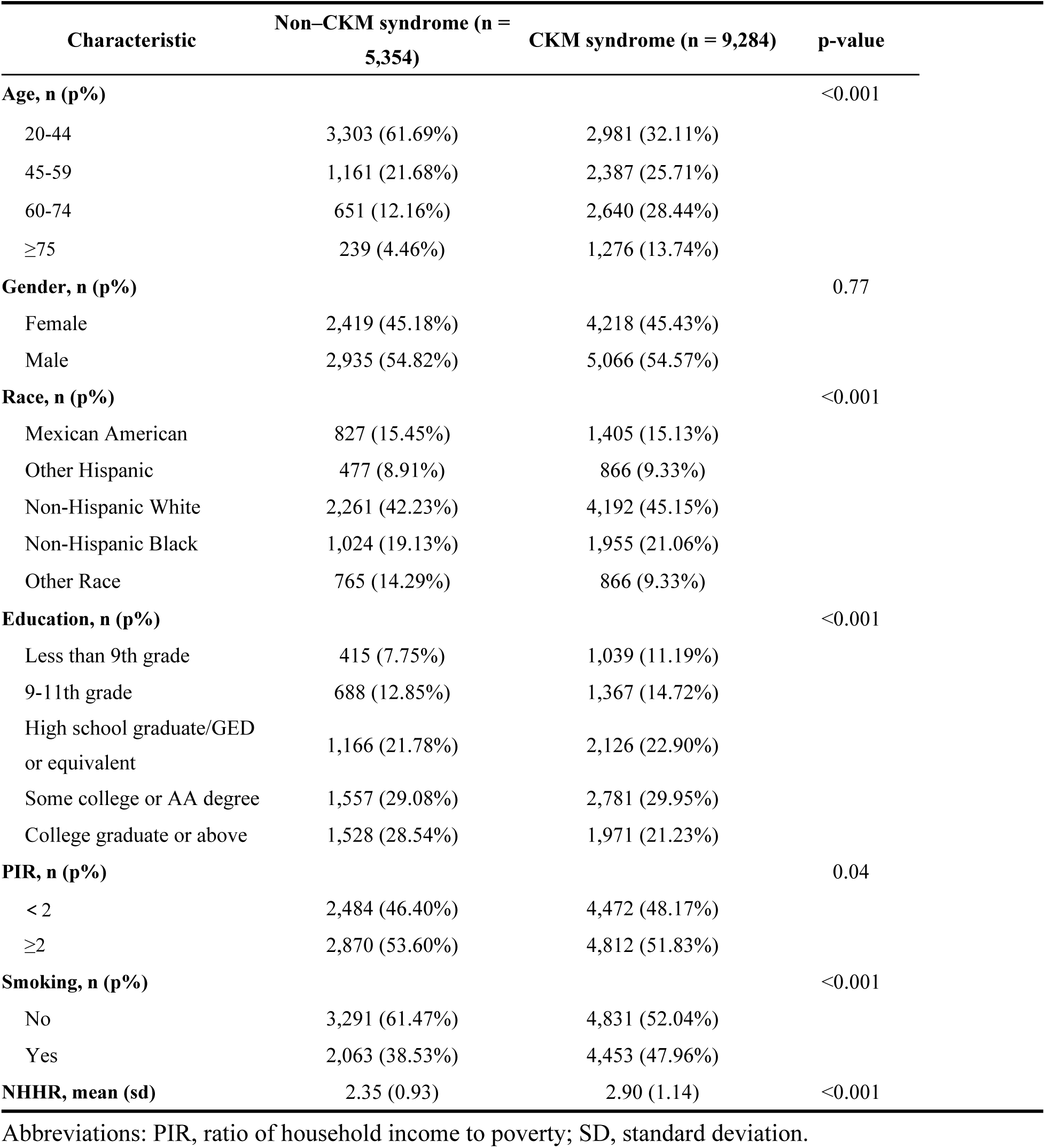
Baseline characteristics by Cardiovascular-Kidney-Metabolic Syndrome.

| Characteristic | Non-CKM syndrome (n = 5,354) | CKM syndrome (n = 9,284) | p-value |
| --- | --- | --- | --- |
| **Age, n (p%)** |  |  | <0.001 |
| 20-44 | 3,303 (61.69%) | 2,981 (32.11%) |  |
| 45-59 | 1,161 (21.68%) | 2,387 (25.71%) |  |
| 60-74 | 651 (12.16%) | 2,640 (28.44%) |  |
| ≥75 | 239 (4.46%) | 1,276 (13.74%) |  |
| **Gender, n (p%)** |  |  | 0.77 |
| Female | 2,419 (45.18%) | 4,218 (45.43%) |  |
| Male | 2,935 (54.82%) | 5,066 (54.57%) |  |
| **Race, n (p%)** |  |  | <0.001 |
| Mexican American | 827 (15.45%) | 1,405 (15.13%) |  |
| Other Hispanic | 477 (8.91%) | 866 (9.33%) |  |
| Non-Hispanic White | 2,261 (42.23%) | 4,192 (45.15%) |  |
| Non-Hispanic Black | 1,024 (19.13%) | 1,955 (21.06%) |  |
| Other Race | 765 (14.29%) | 866 (9.33%) |  |
| **Education, n (p%)** |  |  | <0.001 |
| Less than 9th grade | 415 (7.75%) | 1,039 (11.19%) |  |
| 9-11th grade | 688 (12.85%) | 1,367 (14.72%) |  |
| High school graduate/GED or equivalent | 1,166 (21.78%) | 2,126 (22.90%) |  |
| Some college or AA degree | 1,557 (29.08%) | 2,781 (29.95%) |  |
| College graduate or above | 1,528 (28.54%) | 1,971 (21.23%) |  |
| **PIR, n (p%)** |  |  | 0.04 |
| < 2 | 2,484 (46.40%) | 4,472 (48.17%) |  |
| ≥2 | 2,870 (53.60%) | 4,812 (51.83%) |  |
| **Smoking, n (p%)** |  |  | <0.001 |
| No | 3,291 (61.47%) | 4,831 (52.04%) |  |
| Yes | 2,063 (38.53%) | 4,453 (47.96%) |  |
| **NHHR, mean (sd)** | 2.35 (0.93) | 2.90 (1.14) | <0.001 |

Abbreviations: PIR, ratio of household income to poverty; SD, standard deviation.

### Baseline characteristics of the research participants stratified by NHHR quartiles

The baseline characteristics for categorization based on quartiles of NHHR are shown in **Table 2**. NHHR quartiles were defined as follows: Q1 (1.13–1.75), Q2 (2.00– 2.38), Q3 (2.67–3.18), and Q4 (3.59–4.87). The participants with higher NHHR values tended to be male, Mexican American, or other Hispanic/White participants. In the higher NHHR quartiles, there was a rise in the number of participants in the younger age groups and those who smoked. Additionally, the proportion of participants without CKM syndrome decreased as NHHR increased, whereas the proportion of participants with CKM syndrome gradually increased.

**Table 2.**
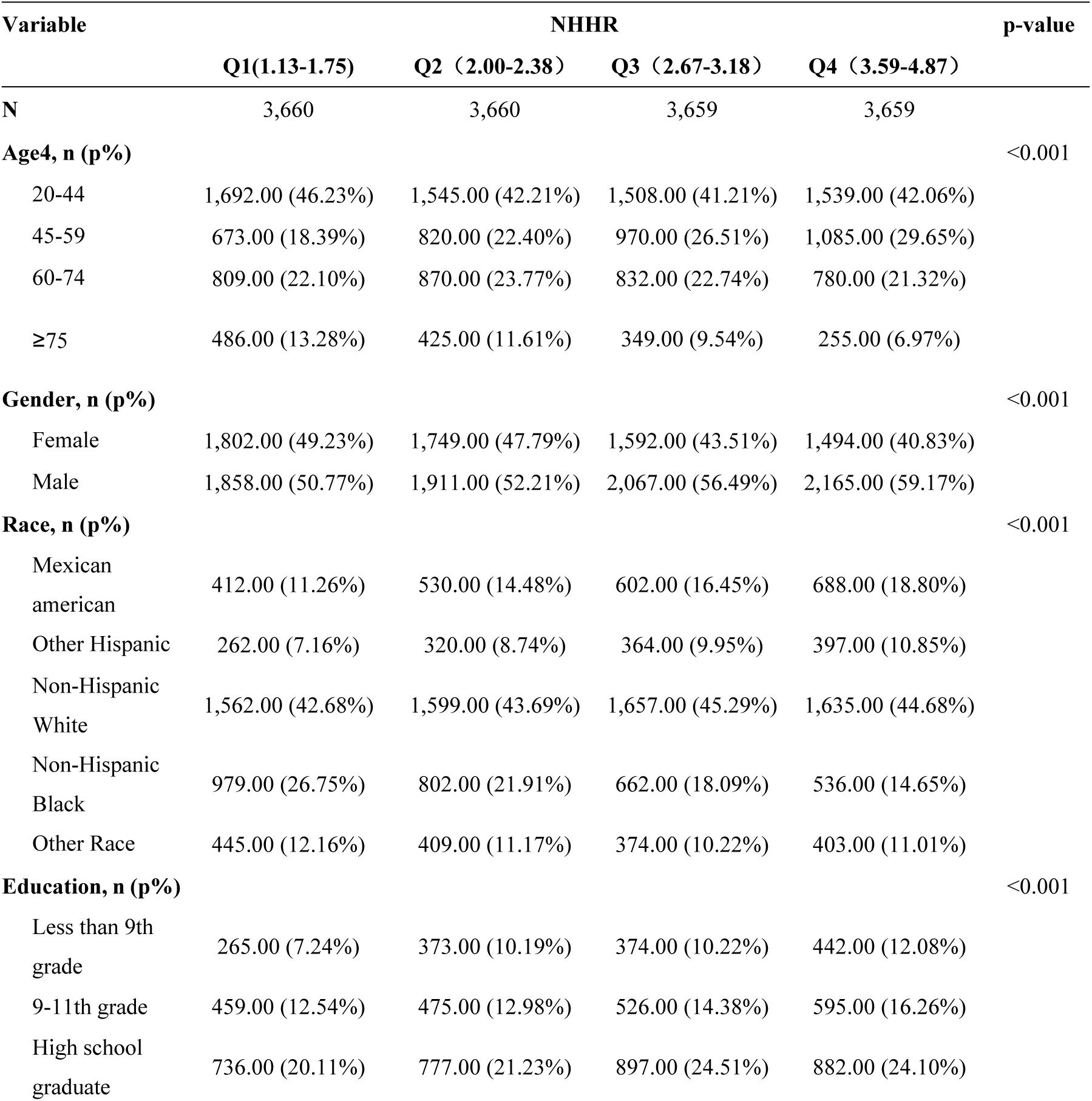
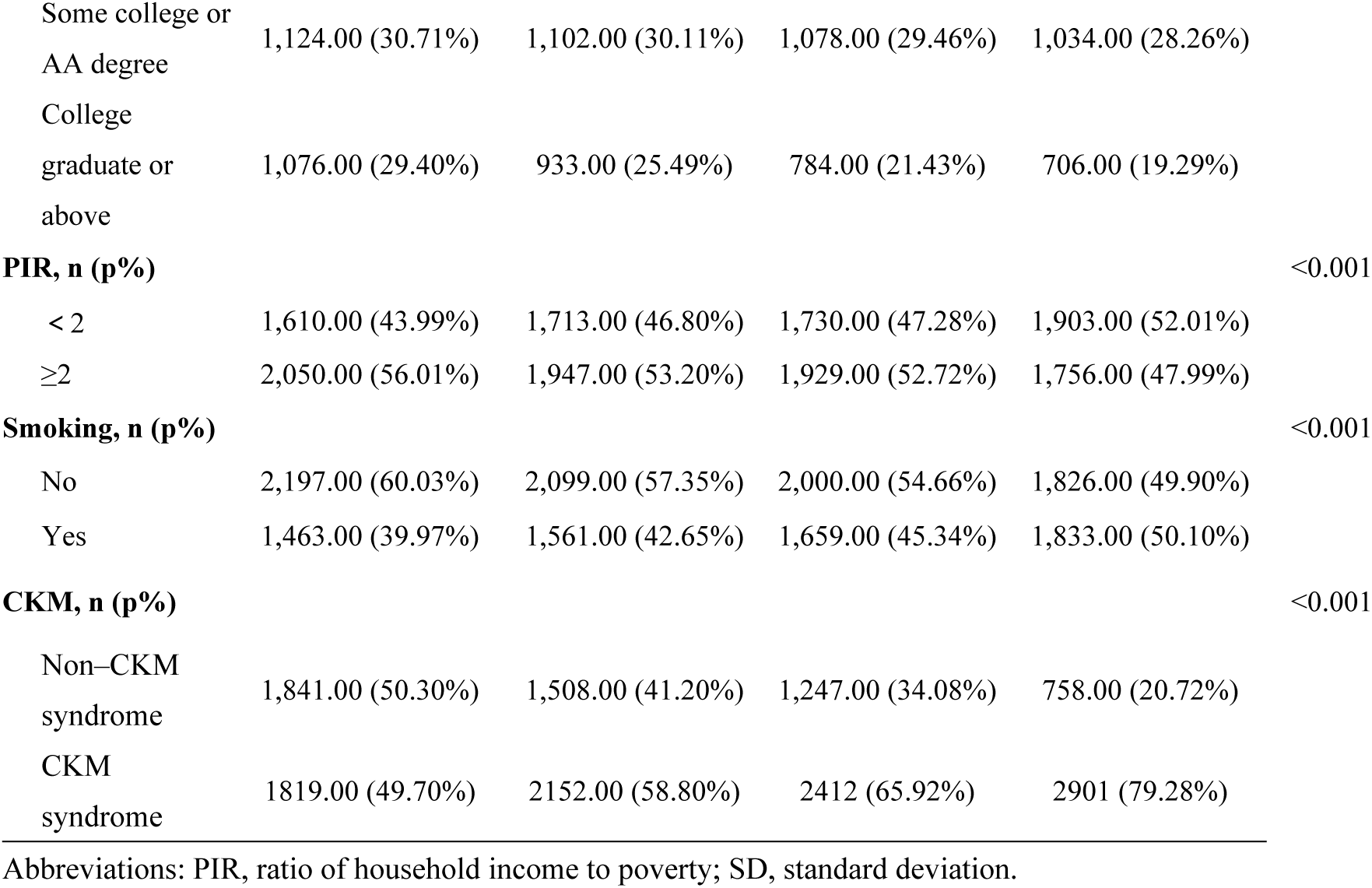
Baseline characteristics according to the NHHR quartiles.

### The association between NHHR and CKM

Figure 2A illustrates the distribution of NHHR levels across stages 0-4 of CKM syndrome. The results revealed that NHHR values in CKM stage 0 were lower than those in CKM stages 1-4, with the median and interquartile ranges at relatively low levels and a narrow distribution range. In contrast, the median NHHR values from CKM stages 1 to 4 in each stage are significantly higher than those in stage 0, with a broader distribution range. NHHR values in CKM stage 2 reach a maximum, with the median being considerably higher than those in the other stages, and the distribution range is the broadest. Further comparison of NHHR distribution in CKM stage 0 with that in the combined stages 1-4 via box-and-line plots (Figure 2B) revealed a significant difference in NHHR values between CKM stage 0 and stages 1-4 (P < 0.001). This trend may suggest that NHHR may play a role in the pathological process of CKM syndrome.

**Figure 2.**
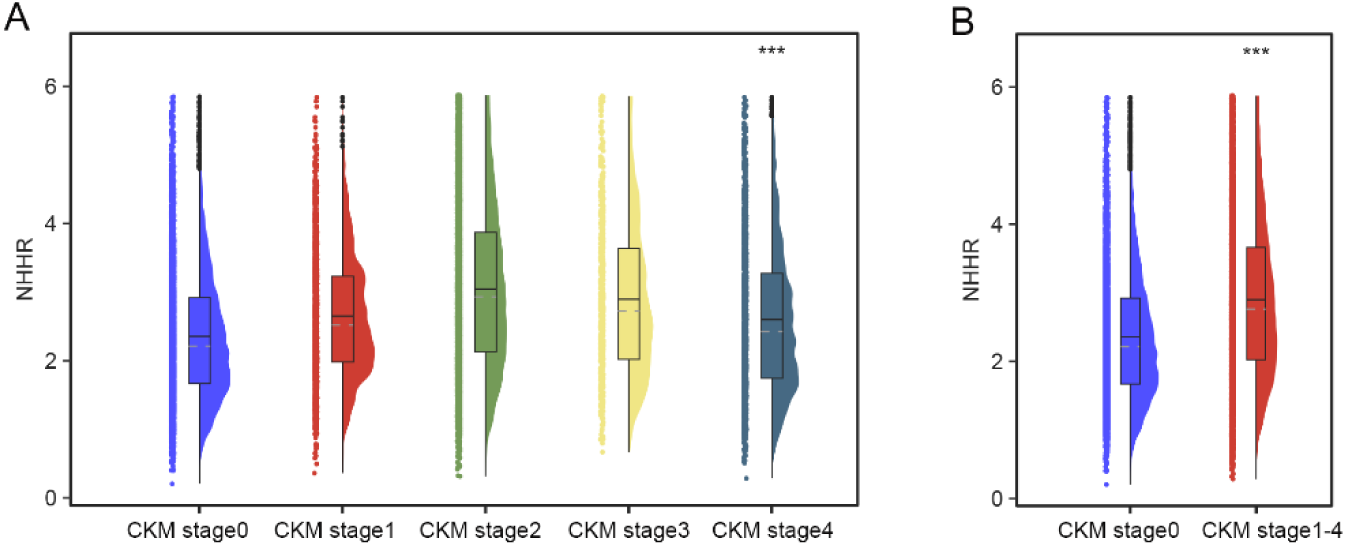
Differences in the distribution of NHHR levels between CKM stages 0-4 (A) and between CKM stage 0 and CKM stages 1-4 (B).

Multivariate logistic regression analyses were conducted to evaluate the connections of NHHR with the occurrence of CKM syndrome and its components, as presented in **Table 3**. The results indicated the unadjusted model showed a positive association of NHHR with CKM (OR = 1.64, 95% CI: 1.59, 1.83), which remained significant in following adjustment for demographic and health-related variables, including age, gender, race, PIR, education level, and smoking (OR = 1.74, 95% CI: 1.70, 1.81). When NHHR was examined by dividing it into quartiles, OR for NHHR quartiles 2, 3, and 4 in Model 3 were 1.47 (95% CI: 1.33, 1.62), 2.10 (95% CI: 1.89, 2.33), and 4.46 (95% CI: 3.99, 4.99), compared with the first quartile (Q1). The potential nonlinear association between NHHR and CKM was illustrated using RCS curves (Figure 3). After adjusting for potential confounders, a significant nonlinear relationship was observed between NHHR and CKM syndrome (nonlinear p < 0.001).

**Figure 3.**
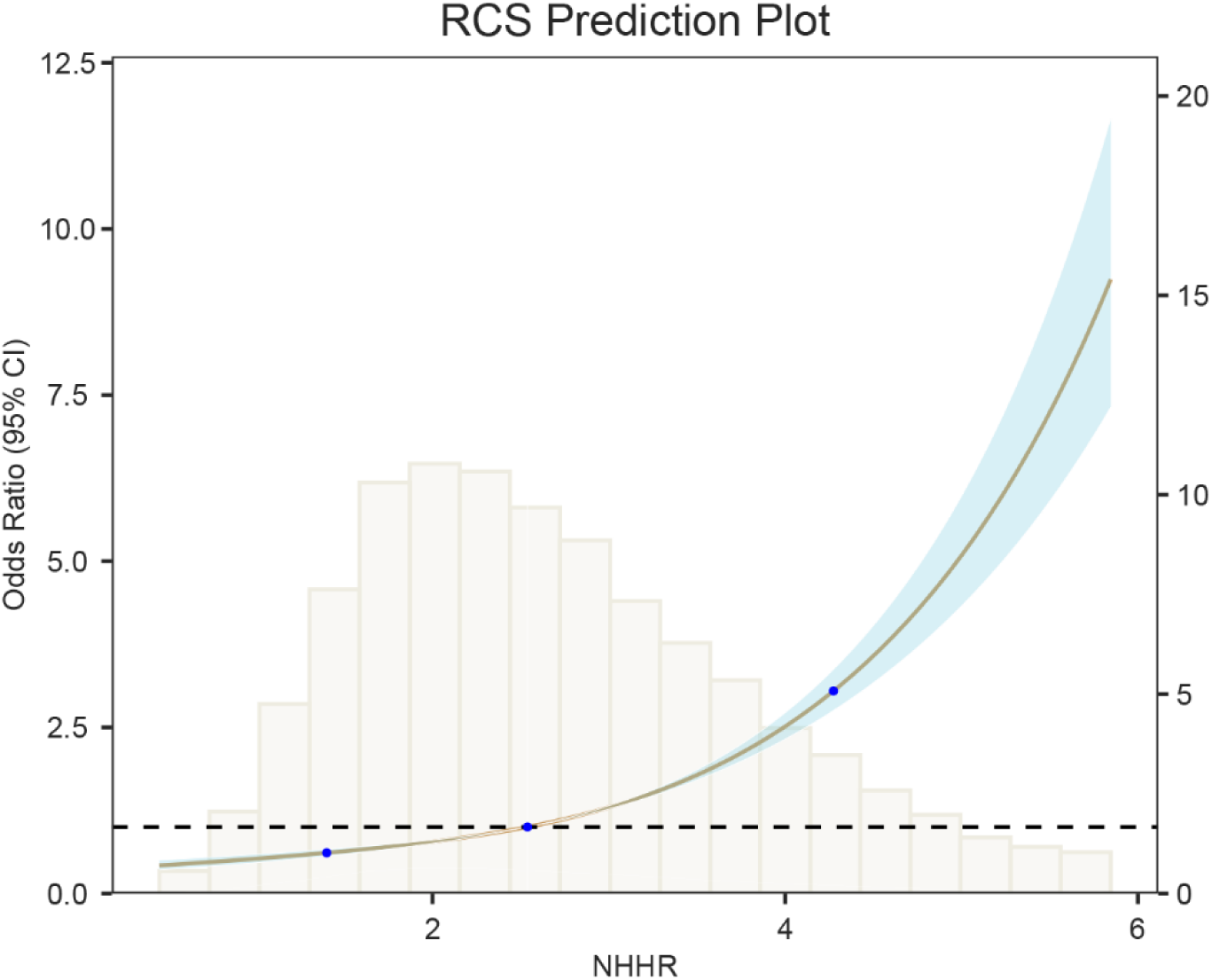
The restricted cubic spline curve of the association between the NHHR and CKM syndrome.

**Table 3.**
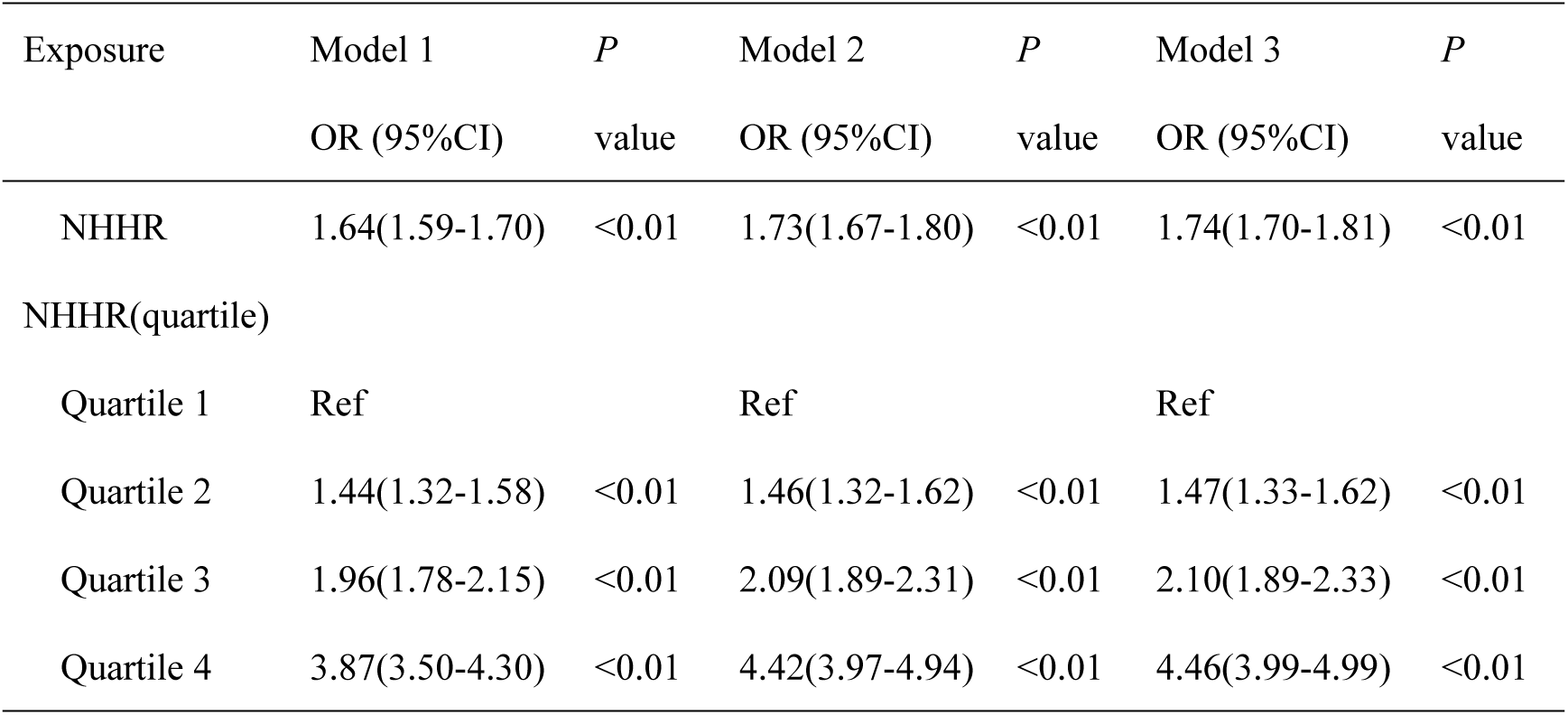
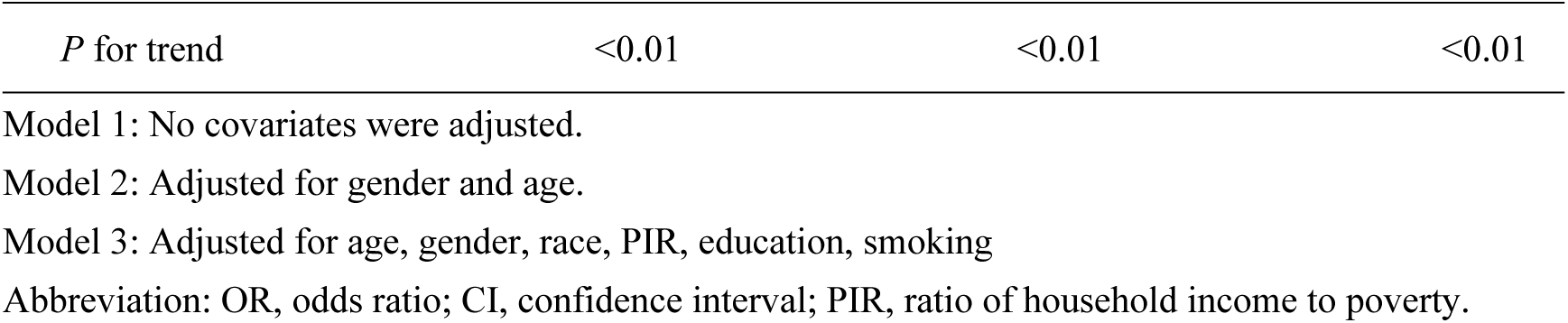
Multivariate regression analysis of NHHR with CKM syndrome.

### Evaluation of the predictive performance of NHHR for CKM stages 1--4

To study the complex connection of the NHHR with CKM stages further, we developed a logistic regression model for CKM stages 1- 4 on the basis of the unadjusted Model 1 (**Table 4**). The results revealed that NHHR was strongly related to the CKM stages 2-4, except for stage 1 (P < 0.01). Among these stages, the highest odds ratio (OR) was found in CKM Stage 2, with an OR of 1.84 (95% CI: 1.77, 1.91), whereas the ORs for CKM Stages 3 and 4 decreased to 1.68 (95% CI: 1.55, 1.82) and 1.28 (95% CI: 1.21, 1.35). The ROC curves (Figure 4) revealed that NHHR had the best ability to discriminate between CKM stage 2 patients, with an AUC of 0.673, followed by CKM stage 3 patients, with an AUC of 0.638.

**Figure 4.**
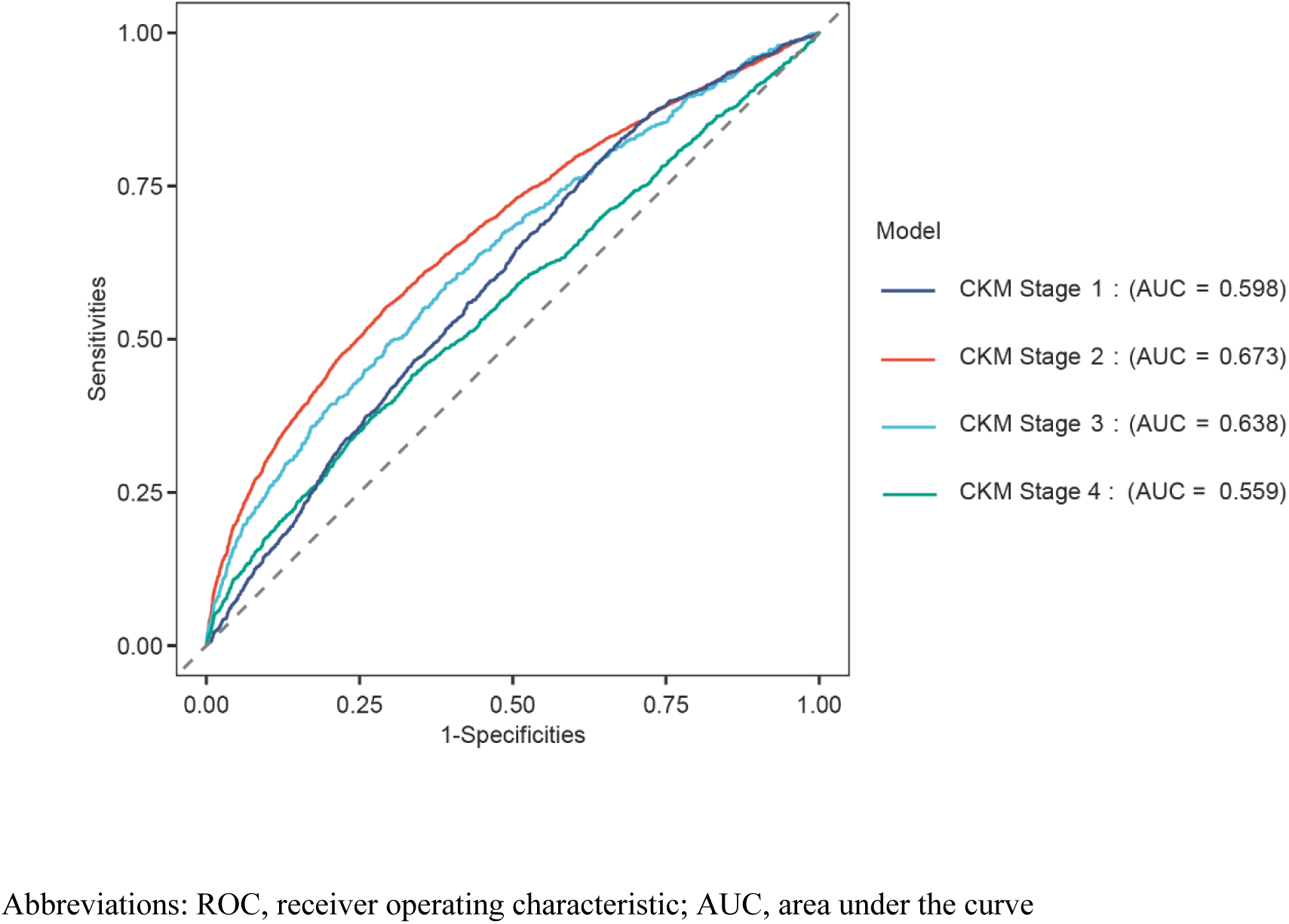
ROC curves comparing the predictive performance of NHHR for CKM stages 0-4.

**Table 4.**
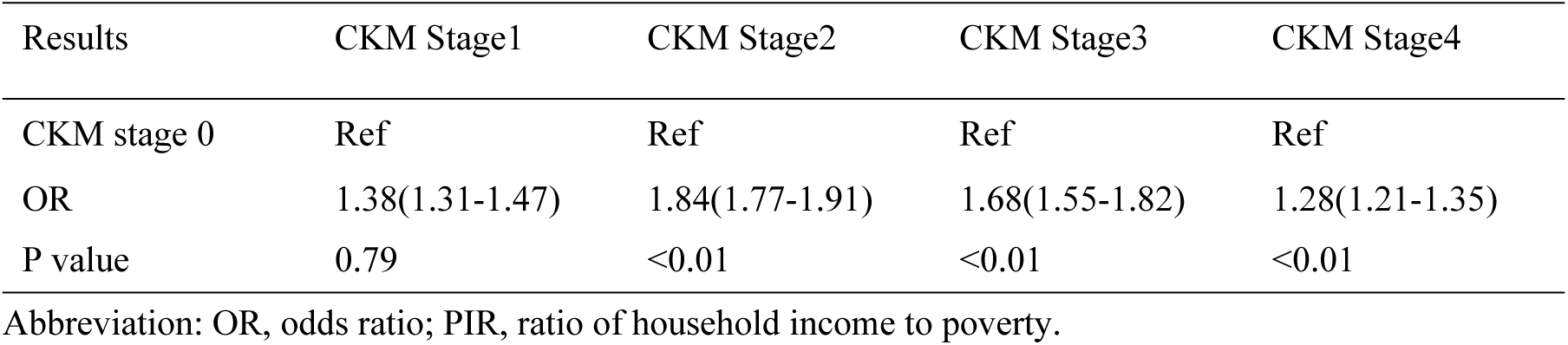
The association between NHHR and CKM stages 1-4 in unadjusted logistic analysis model.

| Results | CKM Stage1 | CKM Stage2 | CKM Stage3 | CKM Stage4 |
| --- | --- | --- | --- | --- |
| CKM stage 0 | Ref | Ref | Ref | Ref |
| OR | 1.38(1.31-1.47) | 1.84(1.77-1.91) | 1.68(1.55-1.82) | 1.28(1.21-1.35) |
| P value | 0.79 | <0.01 | <0.01 | <0.01 |

Abbreviation: OR, odds ratio; PIR, ratio of household income to poverty.

### Subgroup analysis

Subgroup analyses were performed to evaluate the associations of NHHR with CKM syndrome across various subgroups based on age, gender, race, education level, PIR, and smoking (Figure 5). The results indicated that age, race, and education significantly modified the effect of NHHR on CKM syndrome (P < 0.001). In the race subgroup, non-Hispanic Black participants presented the weakest connection between the NHHR and the CKM syndrome (OR = 1.36). In the age subgroup, young and middle-aged participants aged 20-59 presented stronger associations than participants aged 60 or older. In the education level subgroup, the OR increased with higher education levels, suggesting that NHHR was more positively associated with CKM syndrome as the education level increased.

**Figure 5.**
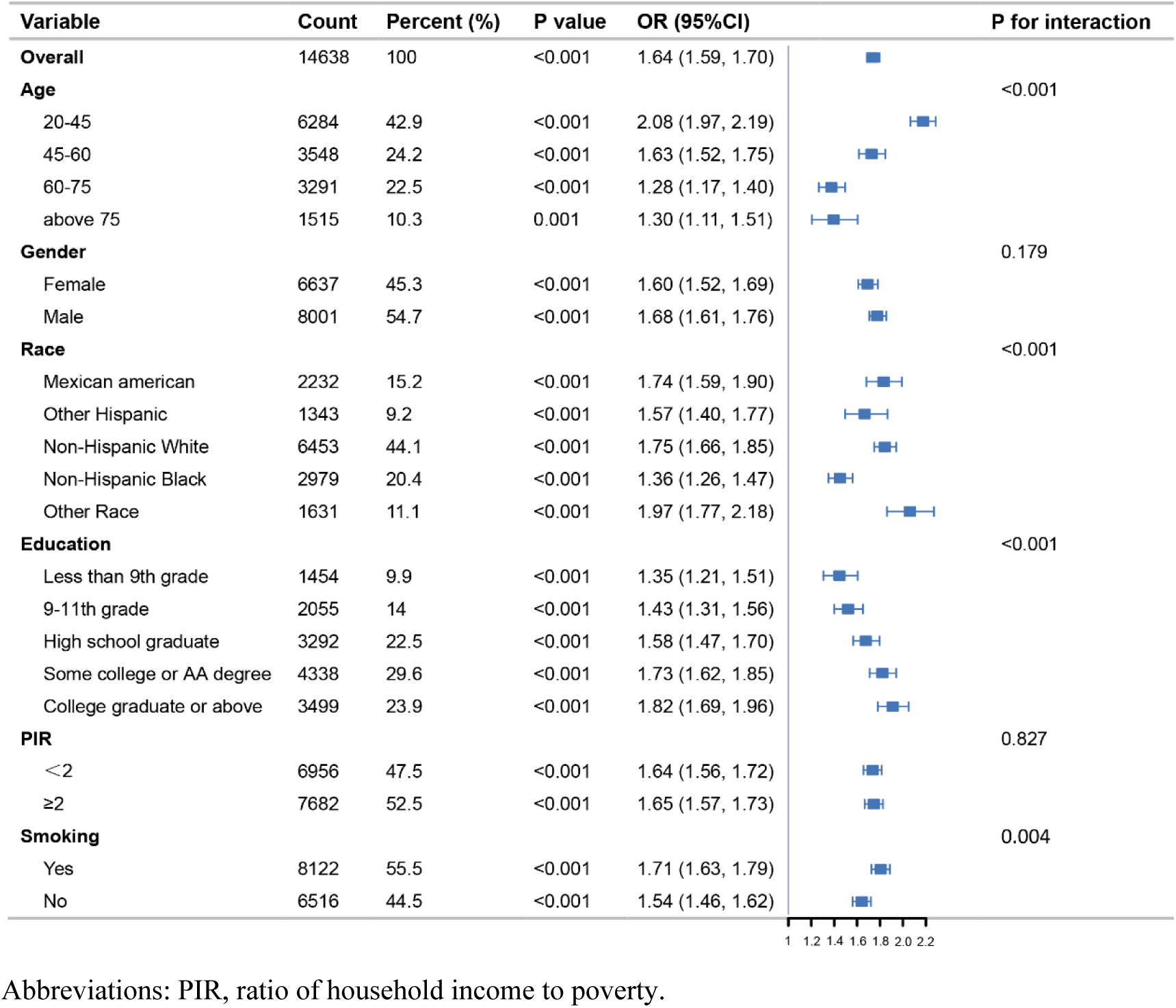
Subgroup analyses of the relationship between NHHR and CKM syndrome.

## Discussion

Data analysis from 14,638 NHANES participants suggested a remarkable association of NHHR with CKM syndrome. Patients with CKM syndrome had significantly higher NHHR levels than participants without CKM syndrome. After adjusting for potential confounders, NHHR remained significantly positively related to the incidence of CKM syndrome. The RCS curves demonstrated a nonlinear relationship between NHHR and CKM syndrome. NHHR showed significant associations and predictive value for CKM stages 2-4, with the strongest associations and predictive efficacy observed for CKM stage 2. Additionally, subgroup analyses and interaction tests revealed that age, race, and education level influence the association of NHHR with CKM syndrome.

In recent years, many studies have shown that NHHR is significantly associated with CKD, CVD, and MetS. Increased NHHR is a risk marker for these diseases and may have an essential predictive role in their occurrence and progression[11–14]. In a study of 83 diabetic patients, Ioana Păunică et al. showed that NHHR showed good sensitivity in identifying high-risk cardiovascular events associated with type 2 diabetes and was superior to traditional lipid indicators [15]. A study covering 451 patients with renal insufficiency undergoing maintenance hemodialysis further found that NHHR is an essential indicator for predicting the occurrence of ischemic stroke and its adverse prognosis, and high NHHR levels are associated with higher stroke incidence and functional impairment[16]. Compared with the above studies, our sample size is more extensive and covers a broader population, improving the results’ reliability. It is worth noting that in the CKM syndrome, a complex and multiple pathological state in which chronic kidney disease, cardiovascular disease, and metabolic syndrome coexist, there are still few systematic studies on the manifestations of NHHR. This study comprehensively explored the changing characteristics of NHHR in CKM syndrome and its association with various stages, providing a more comprehensive basis for clinical diagnosis and treatment.

NHHR integrates information from pro-atherogenic and anti-atherogenic lipoprotein particles, reflecting the balance between lipoproteins. Non-HDL-C encompasses low-density lipoprotein (LDL), very low-density lipoprotein (VLDL), and other triglyceride-containing lipoproteins, such as intermediate-density lipoproteins (IDLs). When non-HDL-C levels are elevated, especially with higher LDL levels, oxidized low-density lipoprotein (ox-LDL) may form through oxidative stress. Ox- LDL activates the inflammatory response, induces dysfunction in vascular endothelial cells, promotes smooth muscle cell migration and proliferation, and forms foam cells that deposit in the arterial wall, contributing to the formation and progression of atherosclerotic plaques[17–19]. In contrast, HDL-C has protective effects, including cholesterol counteraction, anti-inflammatory properties, antioxidant effects, and inhibition of vascular smooth muscle cell proliferation[20]. Elevated NHHR typically indicates elevated non-HDL-C and decreased HDL-C, destabilizing atherosclerotic plaques and triggering thrombosis, remarkably increasing the risk of cardiovascular events[21–24]. NHHR is not only remarkably associated with CVD but also has a critical role in the development and progression of CKD. High non-HDL-C levels disrupt the glomerular filtration barrier by increasing lipid deposition in glomerular mesangial cells and podocytes, leading to cellular dysfunction and apoptosis[25]. Additionally, elevated NHHR induces inflammation and oxidative stress, exacerbating mitochondrial damage and metabolic disorders, which further damage glomeruli and tubules and accelerate CKD progression[26]. Metabolically, NHHR is a comprehensive indicator of lipid metabolism disorders and insulin resistance. Insulin resistance increases lipolysis, increasing the amount of free fatty acids (FFAs) released into the circulation[27]. These FFAs serve as substrates for VLDL synthesis, promoting elevated non-HDL-C levels and disrupting lipid metabolism. Notably, elevated non-HDL-C further exacerbates insulin resistance through lipotoxicity and inflammation, creating a vicious cycle of metabolic imbalance[28]. NHHR plays a key role in impairing the function of the cardiovascular, renal, and metabolic systems through multiple mechanisms, highlighting the importance of exploring its role in the assessment and risk stratification of CKM syndrome.

Our study revealed that NHHR was significantly elevated in patients with CKM syndrome stages 1-4 and had the best predictive performance in patients with CKM stage 2. CKM stage 2 is characterized by the further aggravation of metabolic abnormalities, including hypertriglyceridemia, and related studies have shown that VLDL**s** in non-HDL-C, IDL, **and** chylomicron remnants are highly correlated with nonfasting triglyceride levels[29]. Triglycerides are bound to apolipoprotein B-100 (apoB-100) in the endoplasmic reticulum of hepatocytes through the hepatic metabolism pathway and further loaded into VLDL particles in the Golgi apparatus[30], which results in non-HDL-C elevation; moreover, high triglycerides significantly reduce HDL-C levels through CETP-mediated lipid exchange and increased HDL clearance, ultimately leading to elevated NHHR levels[31]. Notably, this process is regulated by diet, hormones (e.g., insulin), and metabolic disorders[32]. In summary, the hypertriglyceridemia that characterizes CKM stage 2 is more closely associated with elevated NHHR than other stages of CKM, which may be one of the mechanisms explaining the significant elevation of NHHR in CKM stage 2 with the best predictive performance.

Subgroup analysis revealed that the positive connection between NHHR and CKM syndrome was more pronounced in young and middle-aged patients aged 20–59. Consistent with our results, a subanalysis from a multinational consortium database covering a total of 524,444 individuals from Europe, North America, and other countries revealed that non-HDL-C was more strongly related to CVD in individuals younger than 45 years of age than in older individuals (>60 years)[33]. Another study of 9259 individuals in China revealed that the positive connection of NHHR values with CVD was more pronounced in those under 60 years of age[34]. The positive interaction effect between NHHR and age in predicting CKM syndrome may be attributed to several factors. First, a high NHHR in young adults may exacerbate systemic oxidative stress, accelerating the inflammatory response and atherogenesis [35]. Second, long-term exposure to high NHHR status in young people may lead to cumulative cardiovascular and metabolic disorders, and after aging, the vascular elasticity and compensatory capacity of the individual decreases, and the role of NHHR may be masked by other age-related factors (e.g., hypertension and coronary artery calcification)[36,37]. In addition, young people may be more susceptible to lifestyle factors (e.g., a high-fat diet, physical inactivity, smoking, etc.), leading to an increase in risk factors such as obesity, diabetes, and metabolic syndrome [38], which can significantly exacerbate the early onset and progression of CKM syndrome. Therefore, early intervention in young patients with higher NHHR may be an effective strategy to reduce the occurrence and progression of CKM syndrome.

This research has several remarkable advantages. First, this research is the first to systematically assess the relationship between the NHHR and different stages of CKM syndrome, providing a theoretical foundation for its potential use as an early diagnostic tool. Second, this study utilized the extensive 2005-2018 NHANES database, encompassing a substantial sample size and populations from diverse ethnic and socioeconomic backgrounds, enhancing the reliability and generalizability of the findings. However, this research has several limitations. First, owing to its cross-sectional design, this research can reveal an association of NHHR with CKM syndrome but cannot establish a causal relationship. Future studies may use prospective cohort studies to follow-up participants over a long period to clarify the causal relationship between NHHR and CKM syndrome. Second, although a variety of potential confounders were considered (e.g., age, gender, smoking), residual confounding variables, such as genetic factors and dietary habits, may still exist. Detailed information such as genetics, diet, and environment can be further collected to determine possible residual confounding factors and conduct a more precise multifactorial analysis. Additionally, while the NHANES data were collected from a national survey in the United States, the sample predominantly represents the U.S. adult population, which may limit the applicability of the results to other countries or regions. Therefore, future studies can further explore the relationship between NHHR and CKM syndrome in different ethnic and regional populations and optimize the prediction model. At the same time, intervention studies can be conducted to verify whether lowering NHHR levels can effectively improve the prognosis of patients with CKM syndrome and provide more robust evidence for clinical treatment.

## Conclusion

In conclusion, our research confirmed a significant positive connection between NHHR and the risk of CKM syndrome. After adjusting for multiple factors, the risk of CKM syndrome increased by 74% for each unit increase in NHHR. NHHR had a high predictive value for CKM syndrome stage 2, with an AUC of 0.673, and could be used as an important reference indicator for early diagnosis. In clinical practice, NHHR testing can be included in the screening program for people at high risk of CKM syndrome. For patients with elevated NHHR, metabolic, cardiovascular and renal function should be further evaluated, and lifestyle interventions should be adopted, such as a reasonable diet, increased exercise, etc., and lipid-lowering drugs should be given when necessary to reduce the risk of CKM syndrome.

## Nonstandard Abbreviations and Acronyms

AUC: Area under the ROC curve
CI: Confidence interval
CKM: Syndrome Cardiovascular-kidney-metabolic syndrome
CVD: Cardiovascular disease
HDL-C: High-density cholesterol-lipoprotein cholesterol MetS Metabolic syndrome
NHANES: National Health and Nutrition Examination Survey
NHHR: Non-high-density lipoprotein cholesterol to high-density lipoprotein cholesterol ratio
non-HDL-C: Non-high-density lipoprotein cholesterol OR Odds ratio
PIR: Poverty-to-income ratio
RCS: Restricted cubic splines
ROC: Receiver operating characteristic
SD: Standard deviation
TC: Total cholesterol

## Data availability statement

The datasets supporting the findings of this study are publicly available in the National Health and Nutrition Examination Survey (NHANES), which can be accessed at https://www.cdc.gov/nchs/nhanes/index.html.

## Author contributions

JH, Q and J,X developed the study concept and design. JX, T analyzed and interpreted the data. JH, Q and LF, H drafted the manuscript.

### Acknowledgements

We sincerely acknowledge the National Health and Nutrition Examination Survey (NHANES) program for providing valuable data support for this study.

## Declarations Ethics statement

NHANES data collection protocols are approved by the National Center for Health Statistics (NCHS) Research Ethics Review Board, and all participants provided informed consent prior to data collection.

## Consent to statement

All participants provided informed consent to participate.

